# The road to achieving viral hepatitis elimination in Suriname: The progress, the challenges and the goals

**DOI:** 10.1101/2024.01.09.24301061

**Authors:** Giovanni E. Loe-Sack-Sioe, Anfernee Neus, Lycke Woittiez, Terrence Mawie, Frank Coenjaerts, Sigrid Mac Donald – Ottevanger, Stephen Vreden

**Affiliations:** Faculty of Medicine, University of Utrecht; Faculty of Medicine, Anton de Kom University of Suriname; Dept. of Internal Medicine, Academic Hospital Paramaribo, Paramaribo, Suriname; Dept. of Medical Microbiology, Academic Hospital Paramaribo, Paramaribo, Suriname; Dept. of Medical Microbiology, University Medical Center Utrecht, Utrecht, Netherlands; Dept. of Medical Microbiology, Amsterdam University Medical Center, Amsterdam, Netherlands; Foundation for the Advancement of Scientific Research Suriname, Paramaribo, Suriname

**Keywords:** Hepatitis B, Hepatitis C, epidemiology, Suriname, viral hepatitis, elimination, medication, public health

## Abstract

**Background:** Viral hepatitis is one of the leading causes of liver failure, cirrhosis, hepatocellular cancer and mortality worldwide and has long had limited treatment options. Estimates suggest that around 30% of the global population has at one point been infected with HBV, while 3.9% are chronically infected. 1.6 – 2.8% of the world population is estimated to be infected with HCV. With the introduction of the HBV birth-dose vaccines, HBV immunoglobulins and the more recent development of the HCV direct-acting antiviral medication, HBV and HCV infections have now become containable diseases. In 2016, all 194 member states of the World Health Organization endorsed the public-health goal to eradicate viral hepatitis by 2030. Yet, as of 2023, only a small group of countries are on course to achieving this goal. Suriname, a small, multi-ethnic country, located in the north of South America, has an intermediate prevalence of HBV and HCV of 3% and 1% respectively. Previously performed studies, however, show strong ethnic clustering in Suriname.

**Aim:** With this manuscript we dissect the strategy we developed to tackle the hepatitis challenge in Suriname. We explain the importance of establishing more detailed insight into the geographical distribution of high-prevalence areas, as linked to the ethnic differences in the population. We present the screening and contact tracing approaches and end with our current insights on how to proceed toward further prevention and treatment options. Overall, we provide a blueprint towards eliminating viral hepatitis with simple and effective strategies usable in resource-limited countries with an intermediate to high prevalence of viral hepatitis.

## 1. Background

### 1.1 Viral hepatitis and the WHO goals 2030

Viral hepatitis (VH) is the seventh leading cause of mortality worldwide and 81% of these deaths are caused by hepatitis B virus (HBV) or hepatitis C virus (HCV) ^1^. Estimates suggest that over 354 million people are still living with active viral hepatitis and that every 30 seconds at least one person dies from the complications ^2.^ At the World Health Assembly in 2016, countries made the unprecedented commitment to eliminate viral hepatitis globally by 2030 ^2^. Three principal targets were identified: (a) diagnose 90% of people with VH; (b) treat 80% of eligible persons; and (c) reduce incidence and mortality by 90% and 65% respectively ^3^. Since 2016, the number of people receiving treatment for HCV has increased 10-fold ^3^. The WHO has set a goal for 2030 to reduce the prevalence of HBV in children under the age of 5 to 0.1%. However, many countries have failed to meet the targets which were set for 2020. This can be explained by the fact that many countries still lack testing and treatment facilities, together with limited awareness and political commitment. ^2^ To achieve elimination of HBV, women’s and children’s health needs to be prioritized as vertical transmission is the most common transmission route.^1^ Policymakers will have to commit to eliminating viral hepatitis and national policies must be introduced and implemented. Suriname, a multi-ethnic country in South America, with a population of 550,000, of Afro-Surinamese (21.7%), Creole (15.7%), Hindustani (27.4%), Javanese (13.7%), Chinese (1.5%) and Amerindians (3.8%), has also committed to these policies^4^.

### 1.2 HBV and the health burden in Suriname

HBV is one of the most common causes of liver cirrhosis and hepatocellular cancer (HCC) worldwide ^5^. The transmission of HBV has been extensively studied, and it has been found that the primary routes of infection are through blood or other bodily fluids, as well as through congenital transmission ^6, 7^. Factors such as needle-incidents, intravenous-drug use, tattoos, piercings, and hemodialysis have also proven to be common risk factors ^3^. The first HBV vaccine was developed in 1981 and ever since the prevalence of HBV in children under the age of 5 has been declining to an all-time low of 1% in 2019 ^3, 8^. In Suriname, the prevalence of HBV is 3% and since 2003, HBV vaccination in newborns has been implemented ^9^. All pregnant women in Suriname are tested during their antenatal screening. HBV screening during pregnancy helps to identify those whose infants are at risk of perinatal transmission ^10^. Early detection and treatment limits individuals developing chronic HBV infection. Based on current guidelines potentially infected infants should receive immunoglobulins within 12 hours postpartum ^11^. However, since this is often unavailable due to high costs, solely tenofovir disoproxil fumarate is given in the third trimester of pregnancy to prevent vertical transmission as an effective, less expensive alternative. ^12^ Previous research conducted by Mac Donald et al. concluded that the prevalence of HBV in Suriname was substantially higher at 9% within the subgroup formed by people of Javanese descent. Data from Mans et al. revealed that the Javanese community contributes to 40% of all the cases of HCC in the country, which is disproportionally high when considering their contribution to the total population of 14% ^13^. The cause of this overrepresentation is not yet confirmed but could most likely be explained by the high prevalence of HBV within this subgroup. The study by Mac Donald et al. also found strong ethnic clustering of the Indonesian sub-genotype HBV/B3 among Javanese descendants and HBV/A1 sub-genotype among African descendants ^9^. In the same study ethnicity, age, cohabitation and education level were found to be associated with HBsAg positivity ^9^.

### 1.3 HCV and the health burden in Suriname

Transmission of hepatitis-C seems to be restricted to blood contact, being a primary blood borne virus. In Suriname, the prevalence of HCV is an estimated 1% and associated with Javanese ethnicity and cosmetic tattooing. In the Javanese population prevalence was as high as 4% .^14^ Since 2018, treatment with the direct-acting antivirals (DAA’s) has been available in Suriname. As of now, however, widespread use is not possible due to high costs and limited supplies.

### 1.4 The yield of contact tracing

Contact tracing has proven to be a powerful tool in containing the spread of numerous infectious diseases. Yet, contact tracing for HBV and HCV has proven difficult as the infections may have an extensive latency period making it difficult to discover index cases.^15^ This, combined with the excessive costs of testing and limited treatment options in the past has resulted in guidelines advising against systematic implementation of contact tracing for viral hepatitis ^15^. However, with the introduction of the highly effective antiviral medication, HCV has become a curable disease shifting the focus from treatment to detection of new cases. In accordance with the WHO goal to detect 90% of all cases by 2030, the effect of contact tracing needs to be re-emphasized ^2, 15^.

**Objectives**

▪ Gaining insight into the health burden of HBV and HCV in different regions in Suriname.
▪ Detecting the remaining obstacles stagnating the elimination of HBV and HCV.
▪ Developing a viral hepatitis elimination plan specified for a small country with limited funding and treatment options.
▪ Closing the knowledge gap and raising awareness on HBV and HCV in Suriname under general-practitioners and the general population.

## 2. Method

To combat the viral hepatitis epidemic in Suriname, a group of medical professionals initiated the viral hepatitis elimination group of Suriname in 2021. This group developed plans to contain and eliminate chronic viral hepatitis in the country. In July, August and September of 2022 the national viral hepatitis elimination campaign was conducted. This campaign is the first in a series of measures aimed at reducing the transmission, morbidity and mortality of HBV and HCV in Suriname and should provide valuable information on the challenges remaining in the elimination of viral hepatitis. This campaign is based on several pillars consisting of the awareness, detection, prevention and treatment of viral hepatitis in Suriname.

### 2.1 Screening campaign

Twenty regions suspected, based on ethnic variation of inhabitants, to have a diverse prevalence of HBV and HCV were screened ^9, 14^. All participants over the age of 18 willing to get tested were included during the months of July, August and September of 2022. Individuals younger than the age of 18 were only tested as a means of contact tracing. The primary data collected for the screening campaign was non-anonymized, while the data utilized for scientific purposes was subjected to anonymization procedures. Ethical approval was granted by the commission for scientific research in Suriname (CMWO) with document number CMWO30/23. Districts were selected based on earlier collected unpublished data where a high prevalence of HBV and HCV was found in the districts of Commewijne and Nickerie and a lower prevalence in the districts Paramaribo and Wanica. Multiple regions were screened within the districts to gain insight in local clustering. The newly diagnosed positives were identified by screening in these regions via the various local family-practitioner clinics. Screening was performed in a single day per site in which all visiting inhabitants were encouraged to get tested. To further identify the differences in prevalence within regions, multiple villages were visited in all the districts, shown in *Table 3*. HBV and HCV testing was performed via the RDT DetermineHBsAg2 (Abbott, *Illinois, United States of America*) and Oraquick HCV Rapid antibody tests (OraSure, *Pennsylvania, United States of* America) with a reported sensitivity of respectively 99.6% and 98% ^16,17^. Positive index cases were asked to fill out a researcher-led questionnaire regarding their household composition, sexual partner(s), family history and exposure to known risk factors. Defined risk factors included tattoos, piercings, blood-transfusions prior to 1990 for HBV and 1998 for HCV (blood donors were not screened for viral hepatitis before the aforementioned years), hemodialysis and family-members with liver disease or viral hepatitis. Additionally, all newly diagnosed positives were offered laboratory testing, liver elastography and a treatment plan. As HIV co-infection can have an impact on the progression and treatment of viral hepatitis, HIV status was also included in the questionnaire ^18^. As HIV infection was not tested during this campaign, the HIV-status was based on verbally reported information of testing in the past 12 months.

### 2.2 Contact-tracing

The contact-tracing flowchart was developed as a guidance tool into testing the most at-risk contacts. The development of the flowchart was based on the results of a detailed systematic literature search in Embase and Medline for data on the different transmission pathways of HBV and HCV ^19, 20, 21^. The findings were summarized and adjusted to the Surinamese population (*Figures 1 and 2)*.

**Figure 1.**
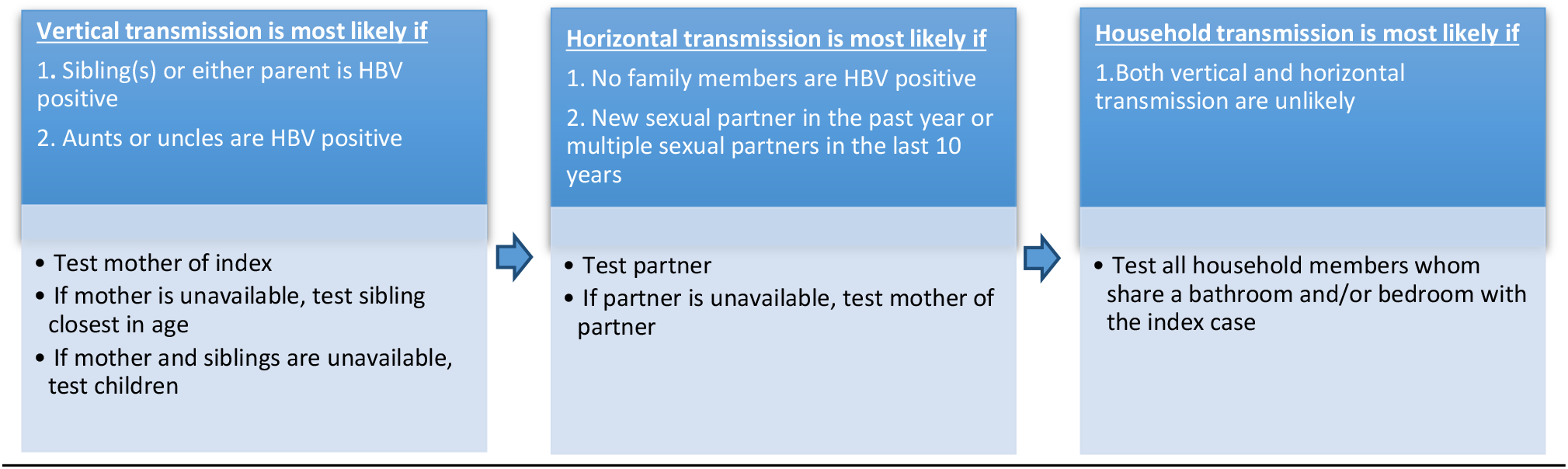
Flowchart for contact tracing implemented for HBV during the screening campaign.

**Figure 2.**
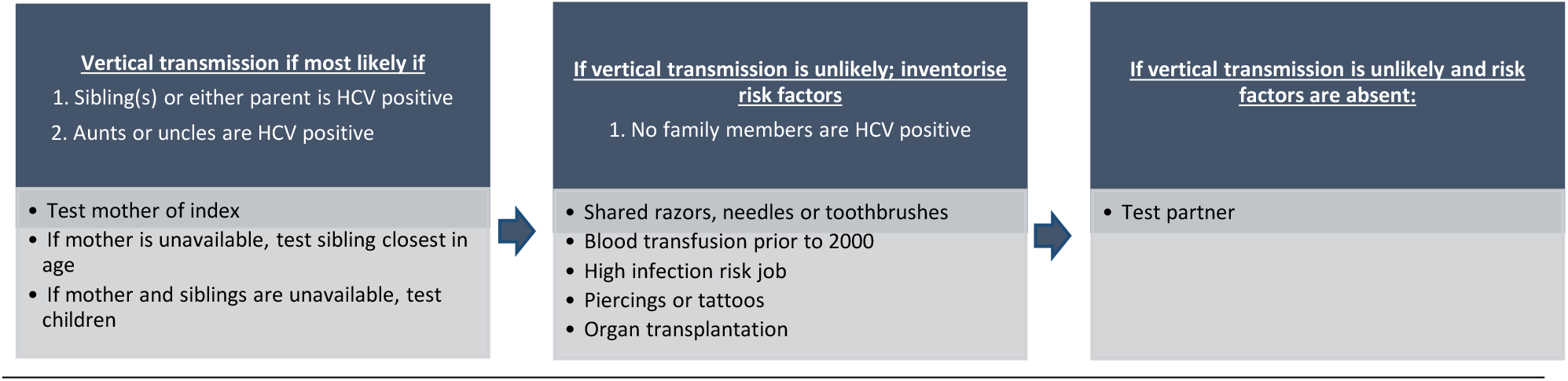
Flowchart for contact tracing implemented for hepatitis-C during the screening campaign.

During this screening campaign, high-risk contacts of newly found positives were notified and invited to get tested. The outcome of contact tracing was defined by determining whether implementation led to the detection of one or more HBV/ HCV cases. Testing of family and household members was done by inviting the contacts to the clinic on the day of screening if they lived or worked nearby or via their family-practitioner if they were not in the vicinity. In this study, all positive cases were informed of the treatment options and the importance of contact tracing. If testing of the at-risk contact was not possible the same day, the index patient was contacted two weeks after diagnosis to inquire whether their high-risk contact(s) had been tested and to report the test outcome.

### 2.3 Treatment

#### HBV

Therapy indications were based on the European Association for the Study of the Liver (EASL) guidelines which are summarized in *Table 1* ^22^. Treatment consisted of tenofovir disoproxil fumarate tablets 245mg, which are provided free of cost by the Ministry of Health of Suriname.

**Table 1:** The different treatment indications based on the combination of viral-load, ALAT liver function test and the degree of fibrosis. These are based on the current EASL guideline on HBV ^22^. NA = not applicable.

| Indications for HBV treatment |  |  |
| --- | --- | --- |
| HBV DNA | ALAT Liver Function Test | Degree of fibrosis |
| Detectable | NA | Cirrhosis |
| > 2000 IU/ml | >1x Upper normal limit | Moderate to Severe fibrosis |
| > 20.000 IU/ml | >2x Upper normal limit | NA |

#### HCV

Treatment indications are based on the EASL guideline for HCV which recommends that all HCV positives with a detectable viral load should be encouraged to receive treatment ^23^. Medication consisted of a combination tablet of sofosbuvir and velpatasvir 400/100mg, taken once daily for a period of twelve weeks. As per the guidelines, three months post-therapy, an additional viral load was performed to assess treatment effectiveness.^23^ Unfortunately, HCV treatment is not provided by the government and if at all, only partially reimbursed by the health insurance companies.

## 3. Results

### 3.1 Screening campaign

A total of 1119 people were screened from the various districts. The baseline characteristics are shown in *Table 2*. Most persons screened lived in the districts of Commewijne and Nickerie, 33.8% and 44.6% respectively *(Figure 3)*.

**Table 2:** Baseline characteristics of all included participants based on gender, pregnancy status, HBV vaccination-status and HIV-status. NA = not applicable.

| Baseline Characteristics | Total | Percentage |
| --- | --- | --- |
| Total number of people tested | 1119 | NA |
| Female/ total number tested | 730/ 1119 | 65.2% |
| Pregnant/ total number tested | 12/ 730 | 1.6% |
| Vaccinated against HBV | 64/ 1119 | 5.7% |
| Known HIV status | 325/ 1119 | 29.0% |
| Known HIV positives | 4 | 1.2% |

**Table 3A:** Detailed information on the HBV and HCV positive cases.

| Characteristics of HBV and HCV positivity |  |  |
| --- | --- | --- |
|  | HBsAg + | Anti-HCV + |
| Total pos./ total number tested | 27 out of 1119 (2.4%) | 66 out of 1119 (5.9%) |
| Female/ Total pos. | 18 out of 27 (66.7%) | 28 out of 66 (42.4%) |
| Pregnant/ Total pos. | 0/ 18 | 0/ 66 |
| Average age | 57.1 years | 54.4 years |

**Table 3B.**
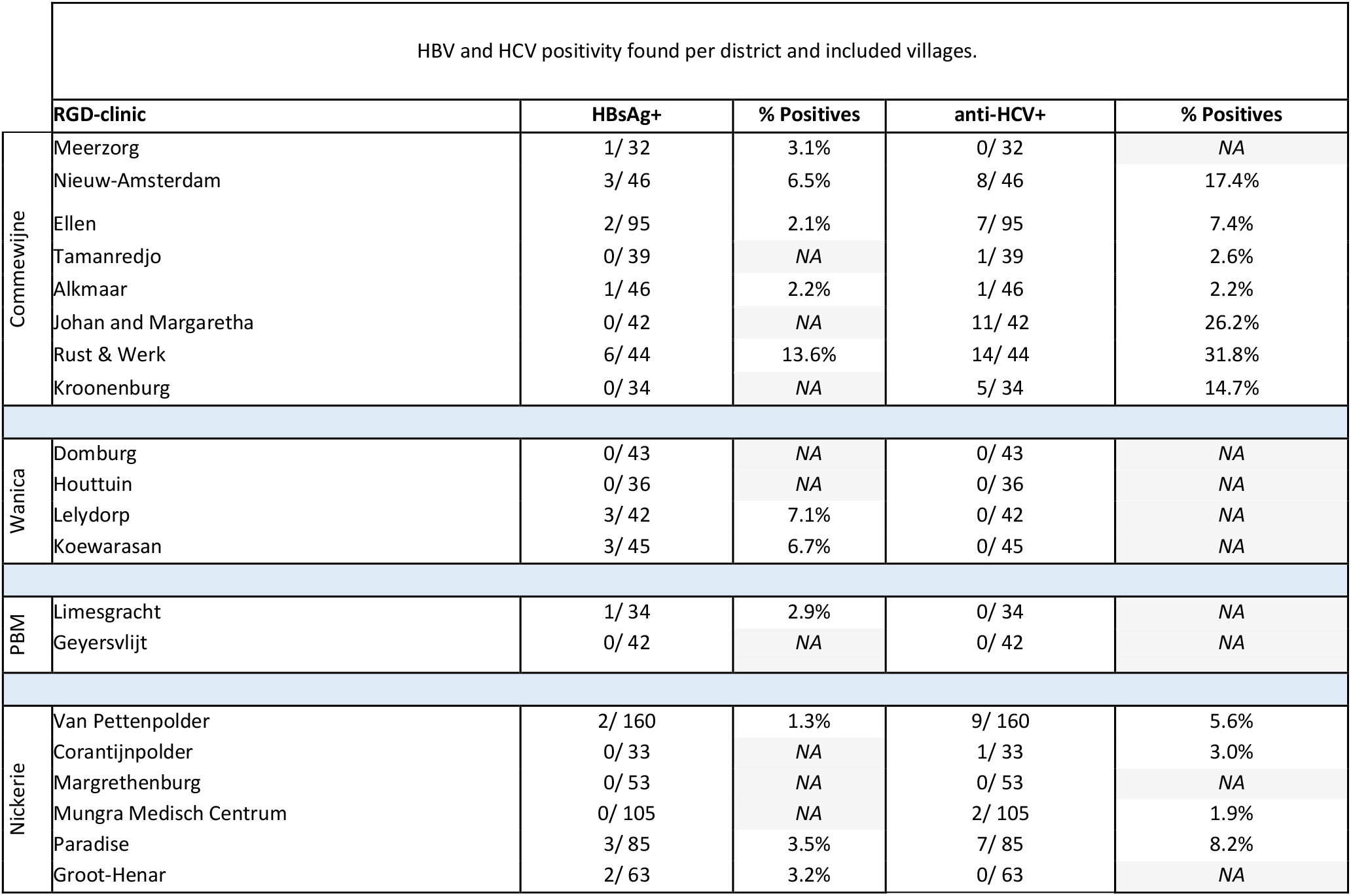
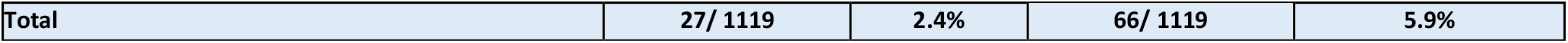
Positive cases in the different villages and their respective districts. The positivity rates per village are also displayed. PBM = Paramaribo. NA = not applicable.

**Figure 3:**
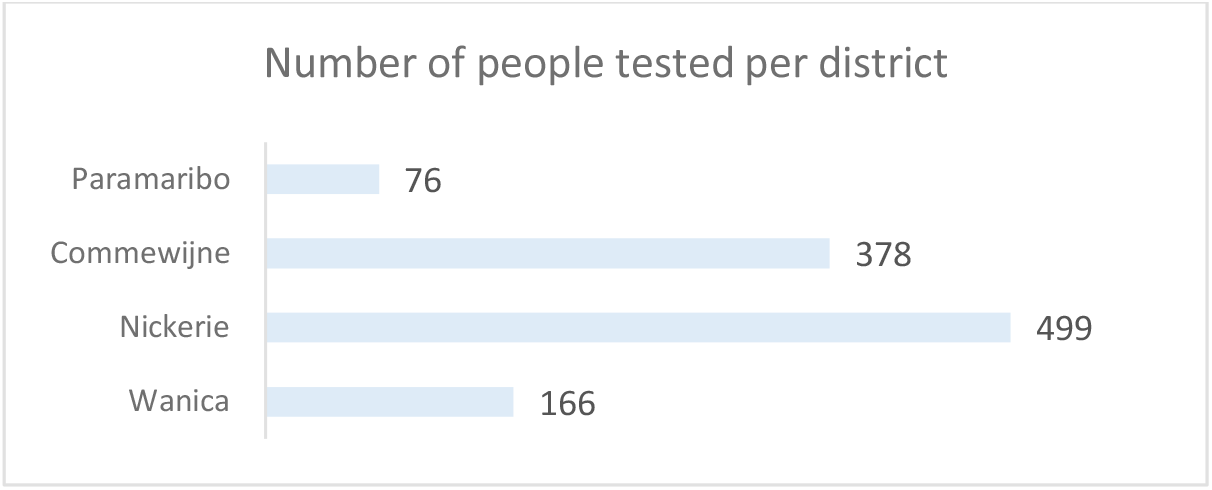
Number of people tested for HBV and HCV in the various included districts.

Out of 1119 people screened, the overall HBV positivity rate was 2.4% and 5.9% for HCV. 42.4% (n= 28) and 66.7% (n = 18) of all positive HCV and HBV cases were females, respectively. None of the HBV or HCV positive women were pregnant. 11.8% had been vaccinated against HBV. Of all the included risk-factors, the presence of a family member with viral hepatitis was the most prevalent. Furthermore, 29% of the participants knew their HIV-status. Of this subgroup 1.2% reported being HIV-positive and receiving treatment. The highest HBV positivity rates were reported in the village of Rust and Werk, whereas the highest positivity rates for HCV were found in Rust and Werk and Johan & Margaretha, both situated in the district of Commewijne. The district of Commewijne had the highest overall seropositivity-rate. Interestingly, all the regions alongside the right bank of the Commewijne-river show a higher HCV positivity-rate when compared to the mainland villages. A detailed overview of the positivity rates per district and village is shown in *Table 3*.

No cases of HCV were discovered in the capital Paramaribo or in the district Wanica (*Figure 4)*. Hepatitis-B positivity was lowest in the district of Nickerie and the capital Paramaribo.

**Figure 4.**
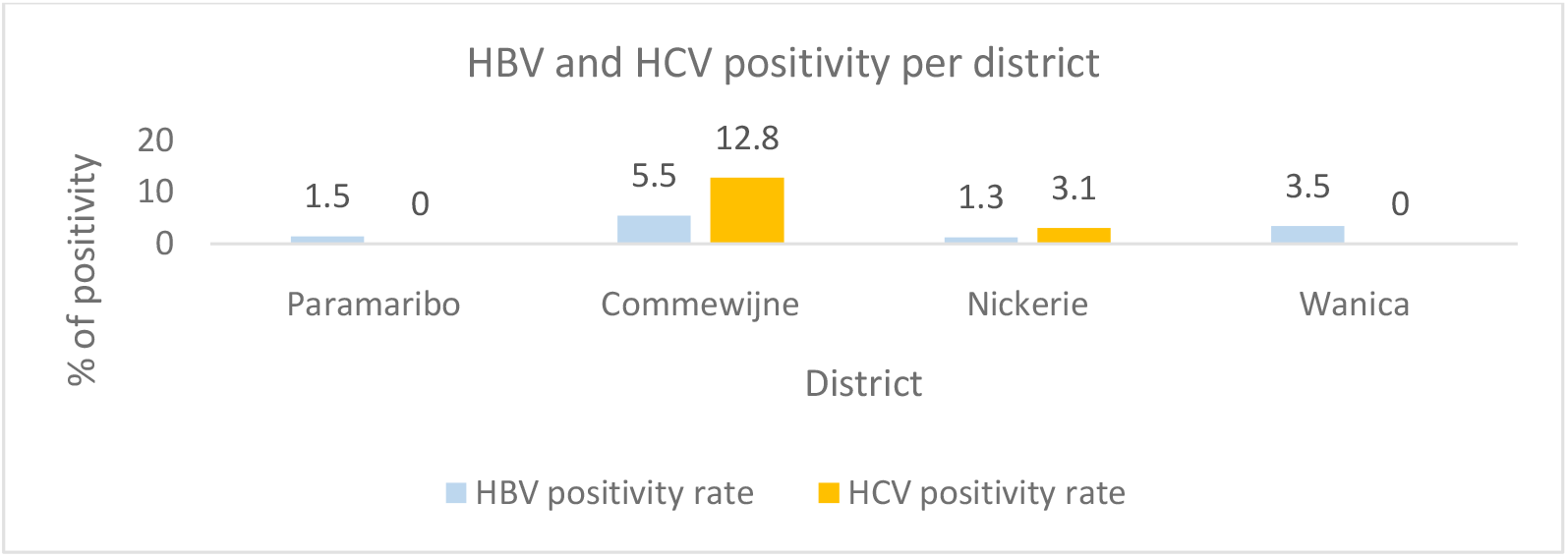
HBV and HCV positivity rates per district.

### 3.2 Contact tracing

In 24% of all HBsAg positive cases, a positive family member was found. 29% of all anti-HCV positives also had a positive family-member *(Figure 5)*. An average of 2 infected family-members per index case were identified. In certain cases, family-members were not available to get tested while the index-patients reported viral hepatitis positivity in a family-member at an earlier point in time. These cases are mentioned separately but are not confirmed by a serological test (*Figure 6)*. 80% of all HBV and 87% of all HCV positive contact cases were based on vertical transmission. In Paramaribo, contact tracing did not lead to the detection of new cases.

**Figure 5:**
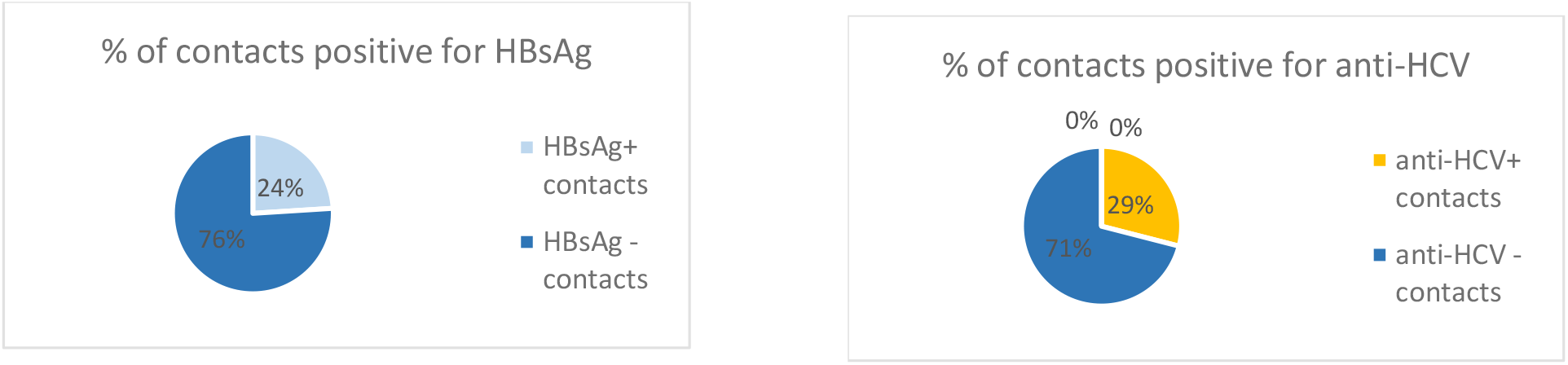
Percentage of serologically confirmed positive HBV (left) and HCV (right) contacts which were discovered as a result of the applied contact tracing in relation to all contacts tested.

**Figure 6:**
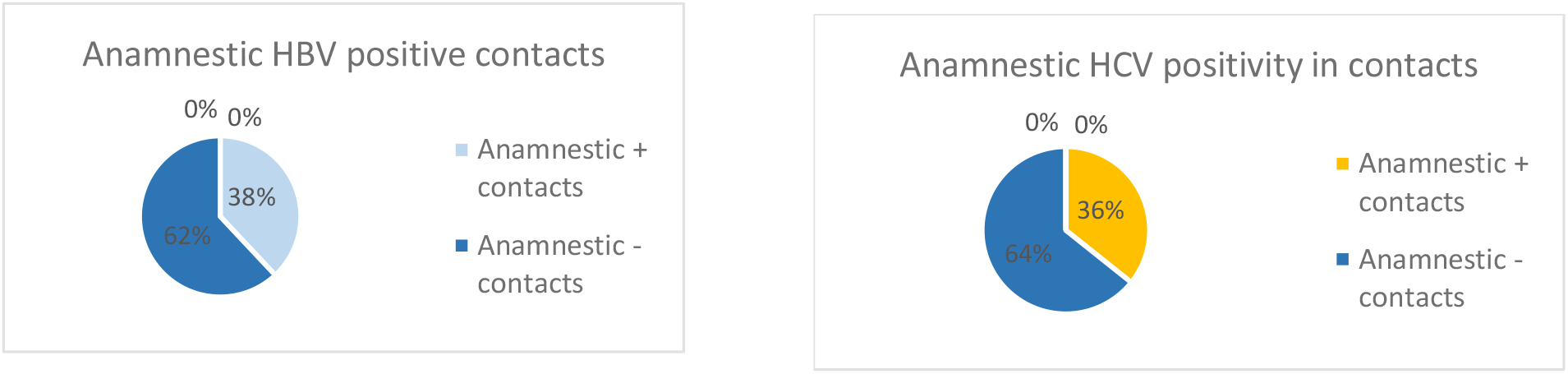
Percentage of verbally stated positive HBV and HCV contacts in relation to all contacts tested.

## 4. Discussion

### 4.1 Screening campaign

The earlier published estimated national prevalence of HBV and HCV is 3% and 1% respectively ^9, 14^. In this study the positivity-rate was 2.4% for HBV (95%CI 0.745% - 4.06%) and 5.9% for HCV (95%CI 1.54% - 10.26%). The higher-than-average HCV seropositivity can be explained by the relatively large sample size from the Commewijne district, where the population is mainly of Javanese descent (HCV prevalence of 4%). Javanese ethnicity has been previously described as a risk factor for viral hepatitis in Suriname by MacDonald et al ^9, 14^. During this campaign these high prevalent regions were consciously chosen to detect new cases as early as possible. Further analysis of the data shows that in villages with a low number of Javanese inhabitants such as Houttuin, Domburg and Corantijnpolder, HCV was hardly detected. Earlier research also showed that HCV genetic subtype 2, which is the only subtype present in Suriname, is directly linked to the subtype found in Indonesia .^14, 24, 25^ This information suggests that the Javanese community in Suriname brought this HCV subvariant into Suriname when they emigrated to the country as contract workers ^24^. Also, the highest positivity-rates are all noted along the right bank of the Commewijne river, in Johan and Margaretha, Rust and Werk, Nieuw Amsterdam and Kroonenburg, which confirms that clustering of infection is present in these villages. The exact reason for clustering remains unknown but might be explained by phylogenetic variations of the HCV virus or by local factors which have not been identified during this study.

### 4.2 Contact tracing

Our data show that implementation of contact tracing leads to an average discovery of two new cases of viral hepatitis per index patient. In 24% of all HBsAg positive cases, a positive contact was found. For anti-HCV this percentage was 29% demonstrating the effectiveness of contact tracing. These results should however be interpretated carefully as screening was performed in the span of a single day per location, and therefore not all contacts could be detected following the contact tracing flowchart. In some instances, index cases came with their entire family in which case all family members were tested without the use of contact tracing. Another interesting aspect is that over 80% of all positive contacts were of mother-child origin, indicating vertical transmission to be an important mode of transmission in the screened population. This furthermore suggests that mother-child transmission may play a larger role than has been acknowledged in previous literature ^19, 20, 21^. A comparable study by Beebeejaun et al. performed in the United Kingdom also found contact tracing to be an effective way to identify, test and vaccinate close contacts of chronic HBV positives, suggesting that contact tracing can be used in a wide variety of settings and countries ^26^. Additionally, Armbruster et al. concludes that screening is favorable in regions with a high positivity rate. As the prevalence reduces, screening becomes less cost-effective thus paving the way for contact tracing to detect new cases. ^27^

### 4.3 Vaccination and treatment

Of all participants included in this study, 11.9% was previously vaccinated against HBV. The low vaccination rate within this study can be explained by the fact that only participants over the age of 18 years old were included for screening. It is expected that a decline in chronic hepatitis-B infections among adults should be noticed in the near future as neonatal vaccination started in 2003. In HBV positive mothers, the international guidelines recommend immunoglobulins for the neonate within the first 12 hours after birth ^11^. However due to the high costs and out of pocket provision, HBV immunoglobulins are often not available to those who need it most.

Treatment with tenofovir disoproxil fumarate has proven to be effective for the treatment of HBV ^11^. To reduce the burden of HBV in Suriname, the Surinamese Ministry of Health is offering medication free of charge which should reduce the progression to liver cirrhosis and hepatocellular cancer in infected people and ultimately reduce HBV-related-mortality. However, as therapy for HBV is not curative and has to be taken for longer periods of time, therapy compliance declines over the course of time ^28, 29^.

The introduction of the DAA’s has resulted in HCV becoming a curable disease ^1,2^. However, as of 2023, none of the insurance companies in Suriname cover the full expenses for this treatment. The costs of approximately 350USD per full treatment is too high given the socio-economic status of most of the Surinamese people. This results in people opting out of therapy and progression of active HCV infection into liver cirrhosis and hepatocellular cancer. Prevention of these complications with the DAA’s is cost-effective on the long term and depicts the need for government and insurance company involvement if elimination of HCV is to be achieved in Suriname ^30^.

### 4.4 Available literature on elimination plans

Over the past decade many first-world countries have developed guidelines to eradicate viral hepatitis. Most of the elimination plans rely on data from first-world countries which propose costly population screening as their cornerstone ^31^. However, in low-and middle-income countries (L(M)IC)’s where testing facilities and finances are limited, such implementation plans are not feasible. Jaquet et al. recommends that simplified testing and treatment methods need to be introduced in low-income settings, lowering the overall costs and simplifying the diagnostic cascade ^31^. The study by Posuwan et al. performed in Thailand recommends that the local and regional health burden of viral hepatitis must be known to successfully eradicate HBV and HCV, for which they recommend solely screening in high-prevalent areas^32^. A similar study by Musabaev et al focuses on the importance of training first-line practitioners in the detection and treatment of viral hepatitis as this could lessen the group which gets lost to follow up ^33^. Based on these studies and on the organization of the national health care plan, the infrastructure and the available finances, a specified elimination plan must be developed with a timeline, elimination goals and the required steps to get there ^31, 32, 33^.

## 5. The remaining challenges in Suriname

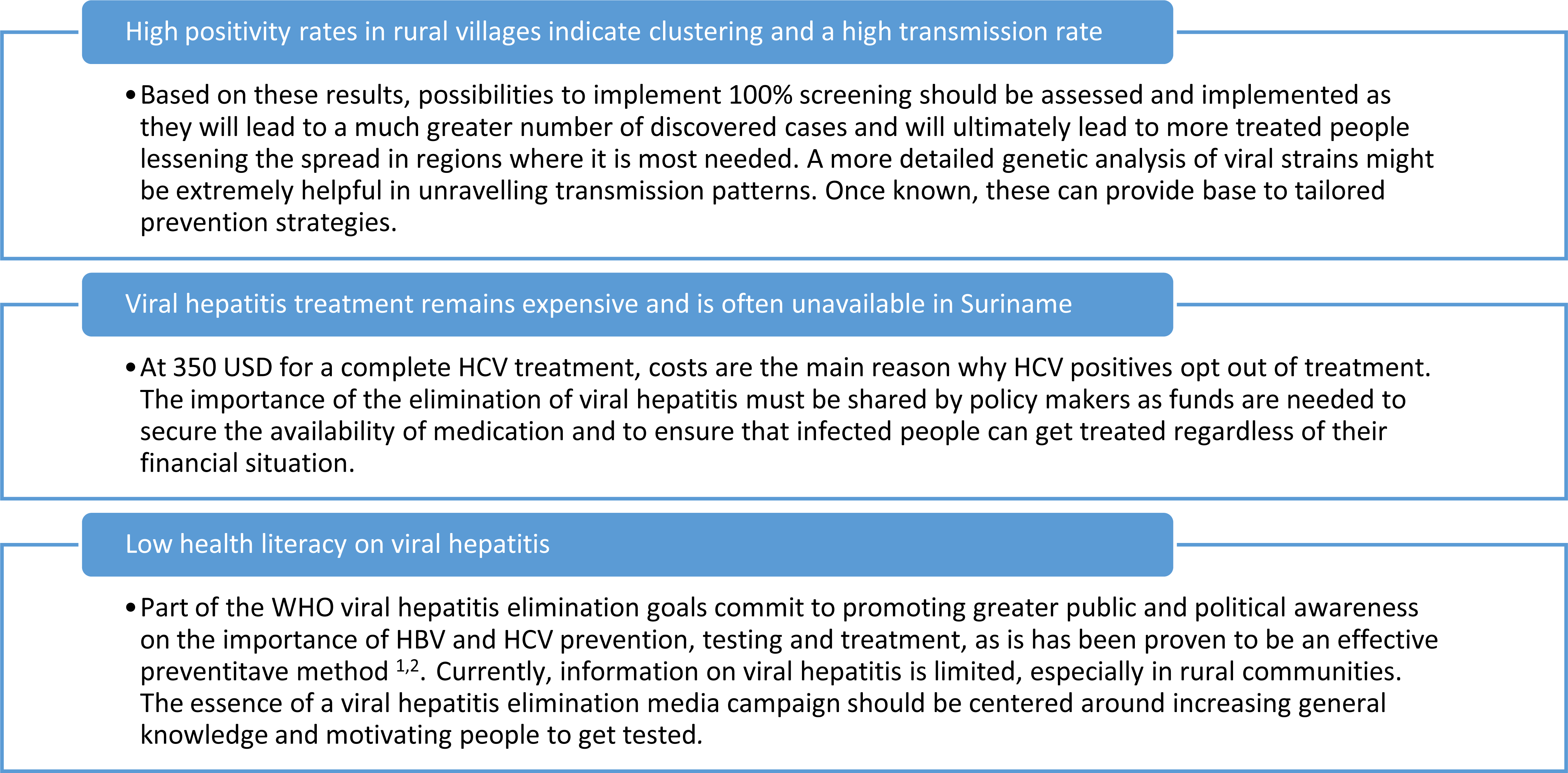

## 6. Conclusion and recommendations

Despite major breakthroughs in the development of treatment options over the past 15 years, communicable diseases such as viral hepatitis still have a major impact on healthcare in many countries ^1, 2, 3^. From the pilot studies we have been able to perform thus far, we can already draw several important conclusions and define additional recommendations. Firstly, in Suriname, Javanese ethnicity is an important risk factor for contracting HBV and HCV ^9, 14^. In rural villages in the district of Commewijne, where the majority of the inhabitants are of Javanese ethnicity, positivity rates approaching 30% were seen underscoring the need for a coordinated plan of action. In these high prevalent regions, though costly, we aim to reach 100% screening as many new cases can be identified relatively easily.

Secondly, contract tracing is a simple, yet effective method to discover new HBV and HCV cases. As HCV infection has become curable, detection of cases will be essential. Thirdly, increasing awareness and up-to-date information under general practitioners will be a key aspect in eliminating viral hepatitis. In line with the recommendation of Musabaev et al, training first-line doctors to motivate, test and treat cases of active viral hepatitis must be prioritized ^33^. The infectious disease specialist should routinely organize courses with the latest developments and information providing guidelines for general practitioners to use.Fourthly, it is of great importance to intensify media campaigns to broaden the public knowledge on viral hepatitis. By providing information on the risk-factors and transmission routes people might be more cautious leading ultimately to less new infections. Finally, all sectors will have to work together to further develop a long-term viral hepatitis elimination plan. For the campaign to be successful the government will have to invest in the implementation of screening and awareness projects on a routine basis. The insurance companies must be made aware of the cost-effectiveness of viral hepatitis treatment coverage ^33, 34, 35^. With all sectors, public-health institutions and professionals working together, viral hepatitis elimination in Suriname is possible in the near future.

## Data Availability

No legal or ethical restrictions to be addressed concerning our data availability. All original data is stored in a online database with is also downloaded to an Excel sheet.

## Acknowledgements

The authors would like to thank the enthusiastic laboratory technicians of the microbiology lab of the Academic Hospital Paramaribo for the laboratory analysis. Moreover, a special thanks to Dr. Irving for designing the database. Lastly, the entire team of fieldworkers should be mentioned for their hard and precise work.

